# Safety, reactogenicity, and immunogenicity of homologous and heterologous prime-boost immunisation with ChAdOx1-nCoV19 and BNT162b2: a prospective cohort study

**DOI:** 10.1101/2021.05.19.21257334

**Authors:** David Hillus, Tatjana Schwarz, Pinkus Tober-Lau, Hana Hastor, Charlotte Thibeault, Stefanie Kasper, Elisa T. Helbig, Lena J. Lippert, Patricia Tscheak, Marie Luisa Schmidt, Johanna Riege, André Solarek, Christof von Kalle, Chantip Dang-Heine, Piotr Kopankiewicz, Norbert Suttorp, Christian Drosten, Harald Bias, Joachim Seybold, EICOV/COVIM Study Group, Claudia Conrad, Doris Steuer, Ute Gläser, Anne-Sophie Sinnigen, Carolin Rubisch, Nadine Olk, Lisbeth Hasler, Angela Sanchez-Rezza, Paolo Kronenberg, Alexandra Horn, Willi Koch, Paula Stubbemann, Julie-Anne Gabelich, Friederike Münn, Julia Tesch, Petra Mackeldanz, Leon Bergfeld, Tobias Bleicker, Jörn Ilmo Beheim-Schwarzbach, Anna Hiller, Sophia Brumhard, Lara Bardtke, Kai Pohl, Daniel Wendisch, Philipp Georg, Denise Treue, Dana Briesemeister, Jenny Schlesinger, Andreas Hetey, Luisa Kegel, Annelie Richter, Ben Al-Rim, Birgit Maeß, Kerstin Behn, Michelle Lysi, Saskia Zvorc, Maria Rönnefarth, Sein Schmidt, Alexander Krannich, Isabelle Schellenberger, Georg Schwanitz, Viktoria Schenkel, Norma Bethke, Claudia Hülso, Sebastian Dieckmann, Christian Peiser, Florian Kurth, Victor Max Corman, Leif Erik Sander

## Abstract

**Objective:** to assess reactogenicity and immunogenicity of heterologous prime-boost immunisations of ChAdOx1-nCoV19 (Vaxzevria, ChAdOx) followed by BNT162b2 (Comirnaty, BNT) compared to homologous BNT/BNT immunisation.

**Design:** prospective, observational cohort study.

**Setting:** unicenter study in a cohort of health care workers at a tertiary care center in Berlin, Germany.

**Participants:** 340 health care workers immunised between 27 December 2020 and 21 May 2021 at Charité - Universitätsmedizin Berlin, Germany

**Main outcome measures:** the main outcomes were reactogenicity assessed on days one, three, five and seven post prime and boost vaccination, and immunogenicity measured by serum SARS-CoV-2 full spike-, spike S1-, and spike RBD-IgG, virus neutralisation capacity, anti-S1-IgG avidity, and T cell reactivity measured by Interferon gamma release assay at 3-4 weeks post prime and boost immunisation.

**Results:** Heterologous ChAdOx/BNT booster vaccination was overall well-tolerated and reactogenicity was largely comparable to homologous BNT/BNT vaccination. Systemic reactions were most frequent after prime immunisation with ChAdOx (86%, 95CI: 79-91), and less frequent after homologous BNT/BNT (65%, 95CI: 56-72), or heterologous ChAdOx/BNT booster vaccination (48%, 95CI: 36-59). Serum antibody responses and T cell reactivity were strongly increased after both homologous and heterologous boost, and immunogenicity was overall robust, and comparable between both regimens in this cohort, with slightly increased S1-IgG avidity and T cell responses following heterologous booster immunisation.

**Conclusions:** Evidence of rare thrombotic events associated with ChAdOx has led to recommendation of a heterologous booster with mRNA vaccines for certain age groups in several European countries, despite a lack of robust safety and immunogenicity data for this vaccine regimen. This interim analysis provides evidence that the currently recommended heterologous ChAdOx/BNT immunisation regimen with 10-12 week vaccine intervals is well tolerated and slightly more immunogenic compared to homologous BNT/BNT vaccination with three week vaccine intervals. Heterologous prime-boost immunisation for COVID-19 may be generally applicable to optimise logistics and improve immunogenicity and to mitigate potential intermittent supply shortages for individual vaccines.

## Introduction

In light of intermittent supply shortages of individual vaccines and evidence of rare, but severe adverse events following vaccination with vector-based vaccines such as ChAdOx1-nCoV19 COVID-19 vaccine (Vaxzevria, AstraZeneca, ChAdOx) [1–4], heterologous prime-boost regimens for COVID-19 vaccines have gained significant interest [5]. Heterologous booster vaccination with an mRNA vaccine following initial immunisation with ChAdOx is now recommended in specific age groups in several countries, including Germany [6], despite limited or absent data on reactogenicity, safety and immunogenicity of this prime-boost regimen in humans.

On January 29, 2021, the German standing committee on vaccination (STIKO) recommended that ChAdOx should only be administered to persons between 18-64 years of age. Consequently, mainly younger persons, including healthcare workers, received ChAdOx while mRNA vaccines (BNT162b2 (BNT) and Moderna mRNA-1273) were prioritized for use in the elderly. In response to reports about rare blood clotting events, including cerebral venous sinus thrombosis associated with ChAdOx vaccination, especially in younger women [2–4], several European countries restricted their recommendations for ChAdOx vaccination to individuals above a certain age limit (e.g. above 60 years in Germany, and 55 years in France) [7]. Heterologous boost immunisation with an mRNA vaccine (BNT or mRNA-1273) was recommended for persons who had already received a first immunisation with ChAdOx, but who are younger than the revised age limit for ChAdOx [7]. In Phase 2/3 trials, both BNT and ChAdOx demonstrated significant reactogenicity, most commonly pain at the injection site, fatigue, headache, chills, and fever, with only a minor fraction of study participants reporting severe reactions [8,9]. A recent interim analysis of reactogenicity data in the Com-COV trial, investigating various heterologous prime-boost regimens of licensed COVID-19 vaccines, reported no serious side effects, but clearly increased reactogenicity following heterologous boost with BNT 28 days after initial vaccination with ChAdOx [10]. In this interim analysis, up to 80% of persons receiving a heterologous prime-boost with ChAdOx/BNT reported fatigue and other systemic reactions, an up to 40-fold increase compared to the respective homologous boost vaccinations [10]. Both BNT and AZ have been shown to elicit robust immune responses with a significant increase following homologous boost vaccination in clinical trials and real world studies [8,9,11–13]. Heterologous prime-boost immunisation has been shown to elicit increased immunogenicity for other vaccines [5,14,15], and early animal experiments suggest increased immunogenicity of boost vaccination with an mRNA vaccine following initial immunization with adeno-vector based COVID-19 vaccines [16]. However, data on immunogenicity of heterologous prime-boost vaccination for COVID-19 in humans is still lacking.

Heterologous ChAdOx/mRNA vaccination has already commenced across Europe, despite a lack of robust immunogenicity and safety data for this combination. No data on immunogenicity and reactogenicity of heterologous versus homologous BNT/ChAdOx vaccination at 10-12 week intervals, as recommended in many countries, is available to date. Here, we report reactogenicity and immunogenicity data of homologous BNT/BNT and heterologous ChAdOx/BNT prime-boost immunisations in a prospective observational cohort study of 340 healthcare workers in Berlin, Germany. We found comparable reactogenicity and robust immunogenicity of homologous and heterologous vaccine regimens.

## Methods

Methodology and study design and assessment of immunogenicity have also been described in detail previously [17].

### Study design

Health care workers receiving routine COVID-19 vaccination were enrolled in the EICOV and COVIM prospective, observational cohort studies conducted at Charité - Universitätsmedizin Berlin, Germany, after written informed consent was obtained. EICOV was approved by the ethics committee (IRB) of Charité - Universitätsmedizin Berlin (EA4/245/20) and COVIM (EudraCT-No. 2021-001512-28) was approved by the Federal Institute for Vaccines and Biomedicines (Paul Ehrlich Institute) and by the Ethics committee of the state of Berlin. Both studies were carried out in accordance with the guidelines of Good Clinical Practice (ICH 1996) and the Declaration of Helsinki.

Study participants either received two doses of BNT three weeks apart or an initial dose of ChAdOx followed by a heterologous boost with BNT 10-12 weeks later, in accordance with the recommendations of the German standing committee on vaccination (STIKO). Baseline data on demographics were collected by questionnaire (eCRF) at enrollment. Blood samples for detection of SARS-CoV-2 specific antibodies and T cell response were collected immediately prior to the first vaccination, and three to four weeks after the first and the second vaccination.

### Assessment of reactogenicity and safety

Participants were asked to fill out electronic questionnaires on reactogenicity, adverse events, medication, and medical visits on days 1, 3, 5, and 7 after the first and second vaccination. In addition, the use of antipyretic medication (NSAID, acetaminophen) before and after vaccination was recorded. We assessed local and systemic reactions to the different vaccines using a modified Food and Drug administration (FDA) toxicity scale [18]. Following the initial assessments, all participants were asked to self-report any systemic symptoms and intake of pain medication through an electronic questionnaire every two weeks. Here, we report on the results of questionnaires collected the first seven days following first and second vaccination.

### Assessment of immunogenicity

Participants with PCR-confirmed infection or detectable anti-nucleocapsid protein (NP) IgG at any time point in the study were excluded from further analysis. A subset of all study participants was selected for immunogenicity analysis based on multivariate matching for sex and age between vaccine groups. Presence of SARS-CoV-2 specific antibodies was investigated using a microarray-based immunoassay including spike (full spike,S1, RBD) and nucleocapsid (N) as antigens in order to discriminate between vaccine-induced antibody response and convalescent SARS-CoV-2 infection (SeraSpot®Anti-SARS-CoV-2 IgG, Seramun Diagnostica GmbH, Heidesee, Germany, [17]). Functional neutralization capacity was investigated using a surrogate SARS-CoV-2 neutralization test (sVNT, cPass, medac GmbH, Wedel, Germany), following the manufacturer’s instructions [17,19]. Maturation of IgG avidity was characterized by a modified anti-SARS-CoV-2 S1 IgG ELISA (anti-SARS-CoV-2 S1 IgG ELISA Kit; Euroimmun Medizinische Labordiagnostika AG, Lübeck, Germany) [17] in 30 randomly selected samples each from the homologous and heterologous boost cohorts who were seroreactive three weeks after prime vaccination. SARS-CoV-2 spike specific T cell responses were measured by an interferon-γ release assay (IGRA; Euroimmun [17]) of S1 peptide stimulated T cells in heparinized whole blood. IFN-γ was measured by ELISA and an arbitrary unit was displayed subtracting the blank optical density (OD) 450/620nm from S1-peptide-pool stimulated samples.

### Statistics

Data are presented as median and interquartile range, unless stated otherwise. Statistical analysis was performed using GraphPad PRISM statistics version 27.0 (IBM Deutschland, Ehningen, Germany). Group comparisons were performed in a univariate analysis using Fisher’s exact test or nonparametric Kruskal-Wallis test with Dunn’s multiple comparisons test. All 95% confidence intervals were calculated according to the Wilson and Brown method [20]. P-values <0.05 were considered statistically significant.

## Results

From December 27, 2020, to March 30, 2021, a total of 340 healthcare workers were enrolled at Charité - Universitätsmedizin Berlin, Germany. Twenty-six participants had either a positive PCR result for SARS-CoV-2 during the study or a detectable anti-spike IgG or anti-N IgG antibody response at baseline or follow up (N), and were therefore excluded from further analysis. Eight participants opted for a homologous ChAdOx/ChAdOx booster immunisation and were only included for reactogenicity analysis of prime immunisation. Baseline characteristics of the study population, an overview of the vaccine groups and respective study sub-cohorts for reactogenicity and immunogenicity analyses are given in **Table 1**.

**Table 1:** Baseline characteristics and schedule of BNT/BNT homologous prime and boost and ChAdOx/BNT heterologous prime and boost study participants. ^1^BNT: BNT162b2 mRNA COVID-19 vaccine, ^2^ChAdOx: ChAdOx1-nCoV19 COVID-19 vaccine, IQR: interquartile range

| Vaccine group | BNT/BNT<sup>1</sup> homologous boost |  | ChAdOx<sup>2</sup>/BNT heterologous boost |  |
| --- | --- | --- | --- | --- |
| Prime to boost interval , median days (IQR) | 21 (21-21) |  | 71 ( 70-73) |  |
| Prime and boost vaccination | 1<sup>st</sup> BNT, n=179 | 1<sup>st</sup> BNT / 2<sup>nd</sup> BNT n=189 | 1<sup>st</sup> ChAdOx n=151 | 1<sup>st</sup> ChAdOx / 2<sup>nd</sup> BNT n=110 |
| **Reactogenicity data, n** | 178 | 159 | 148 | 99 |
| Age, median years (IQR) | 34 (29-44) | 34 ( 29-43) | 35 (28-47) | 37 (29-51) |
| Female, n (%) | 98 (55.0%) | 87 (54.7%) | 101 (68.2%) | 77 (77.8%) |
| **Serology data measured, n** | 94 | 101 | 57 | 61 |
| Δvaccination to sampling, median days (IQR) | 21 (21-21) | 28 (27-30.5) | 26 (22-28) | 21 (121-21) |
| Age, median years (IQR) | 35 (30.75-48) | 35 (30.5-47.5) | 38 (31-52.5) | 38 (30.5-51.5) |
| Female, n (%) | 66 (70.2%) | 73 (72.3%) | 46 (80.7%) | 47 (77.1%) |

### Reactogenicity

All vaccinations were associated with a relatively high frequency of local reactions, most commonly pain and tenderness. Local reactions were usually mild or moderate (**Fig. 1A, B, Table S1**). No major differences were observed in the frequency or severity of local reactions after either of the prime or boost immunisations, with the exception of a slightly higher frequency of local reactions after heterologous ChAdOx/BNT booster vaccination in comparison to homologous BNT/BNT booster vaccination (**Fig. 1A, B, Table S1**). In contrast, notable differences were reported for systemic reactions. These were most frequently reported following prime immunisation with ChAdOx (86.49%, 95%CI: 80.05-91.08), and after homologous BNT/BNT booster immunisation (64.78%, 95%CI: 57.09-71.78), whereas only 51.52% (95%CI: 41.80-61.12) of participants after the heterologous BNT booster (ChAdOx/BNT) vaccination, and 38.76% (95%CI: 31.92-46.09) of participants after the first immunisation with BNT reported systemic reactions (**Fig. 1C**). Severe systemic symptoms, including fatigue, myalgia, headache, feverishness or chills, and fever >38°C, were reported more frequently following ChAdOx prime immunisation and homologous BNT/BNT booster immunisation compared to heterologous ChAdOx/BNT booster vaccination (**Fig. 1D, Table S1**).

**Fig. 1.**
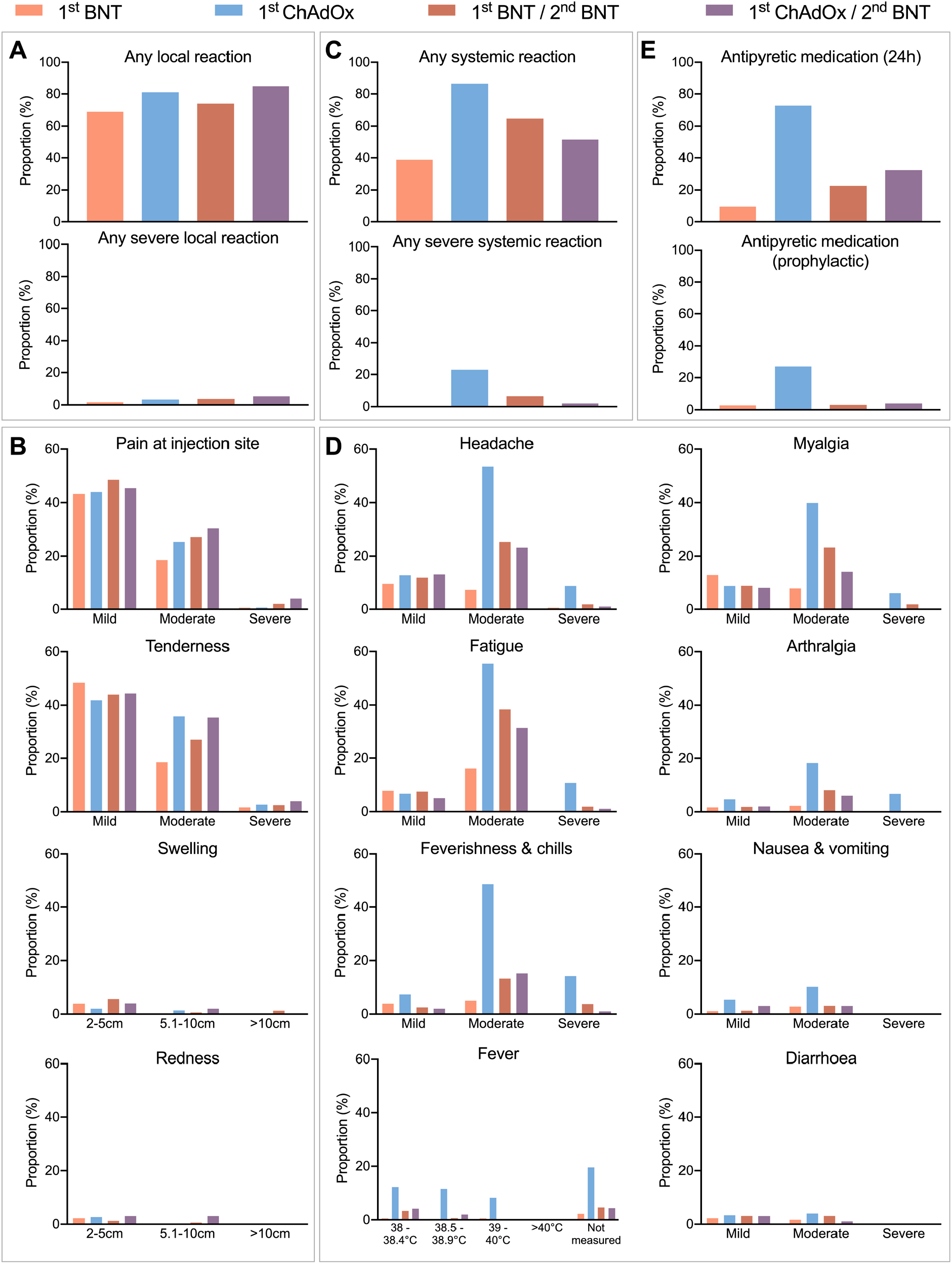
Local and systemic reactogenicity of BNT or ChAdOx prime immunisations and homologous or heterologous boosting until day seven after vaccination. (**A, B**) Proportion of participants reporting any local reaction (A), and indicated local reactions grouped by severity (B). (**C, D**) Proportion of participants reporting any systemic reaction (C), and indicated systemic symptoms grouped by severity (D). (**E**) Proportion of participants reporting intake of antipyretic medication within 24 hours after vaccination (top) and prophylactic intake of antipyretic medication (bottom). BNT: BNT162b2 / Comirnaty, ChAdOx: ChAdOx1-nCoV19 / Vaxzevria. Definition of severity according to modified Food and Drug administration (FDA) criteria [18]: mild: does not interfere with daily activities, moderate: interferes with daily activities, severe: daily activities no longer feasible.

No potentially life-threatening reactions were reported after any of the vaccine regimens in this study. Intake of antipyretic medication was reported most frequently in conjunction with the first ChAdOx immunisation (**Fig. 1E**). Within 24 hours after the first vaccination with ChAdOx, 72.97% (95%CI: 65.30-79.48) of participants reported antipyretic medication, which was markedly lower following heterologous ChAdOx/BNT boost (32.32%, 95%CI: 23.92-42.05), homologous BNT/BNT boost (22.64%, 95%CI: 16.83-29.75), and after prime immunisation with BNT (9.55%, 95%CI: 6.05-14.76) (**Fig. 1E**). The proportion of participants who reported prophylactic antipyretic medication was highest in the ChAdOx prime immunisation group (27.03%, 95%CI: 20.52-34.70), and distinctly lower in all other groups (1.BNT: 2.81% (95%CI: 1.21-6.41), 1.ChAdOx/2.BNT: 4.04% (95%CI: 1.58-9.93), 1.BNT/2.BNT: 3.14% (95%CI: 1.35-7.15). Thus, prophylactic intake of antipyretics cannot explain differences in adverse reactions between ChAdOx/BNT boost vaccination compared to ChAdOx prime vaccination.

The majority of vaccine reactions were reported on day one and three after vaccination and receded by day seven (**Fig. 2**).

**Figure 2:**
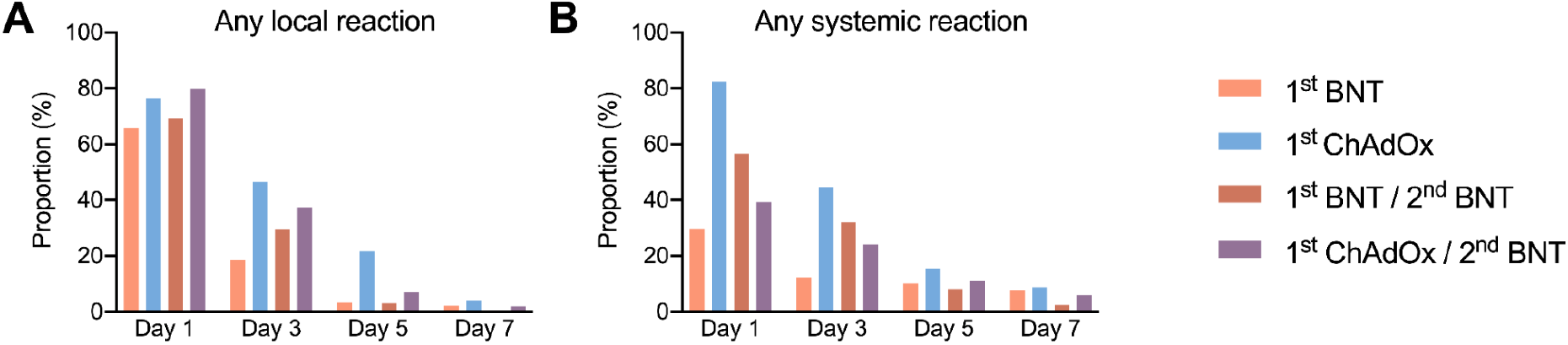
Reactogenicity of BNT or ChAdOx prime immunisation and homologous or heterologous booster vaccination reported until day seven after vaccination. (**A**) Local reactions (any severity) reported by day, over the course of seven days. (**B**) Systemic reactions (any severity) reported by day, over the course of seven days.

### Immunogenicity

Three weeks after prime immunisation with BNT, 63/94 (67.02%, 95%CI: 57.01-75.69) participants were reactive for anti-SARS-CoV-2-S1 (S1) IgG compared to only 16/57 (28.07%, 95%CI:18.08-40.43, p<0.0001) participants after ChAdOx prime immunisation (**Fig. 3A**). The proportion of S1 reactivity increased to 100/101 (99.01%, 95%CI: 94.60-99.95) three weeks after homologous BNT/BNT boost immunisation, and to 61/61 (100.00%, 95%CI:94.08-100.00) three weeks after heterologous ChAdOx/BNT boost immunisation (**Fig. 3A**). Compared to BNT immunised participants, ChAdOx immunised participants had significantly lower anti-S1 IgG levels three weeks after prime immunisation (median 2.08 S/Co, IQR:1.45-3.04 vs 0.52 S/Co, IQR: 0.28-1.00, p=0.02, **Fig. 3A**). Levels of anti-RBD IgG (median 1.28 S/Co, IQR: 0.57-2.16 vs 2.84 S/Co, IQR: 2.02-4.06, p=0.14) and anti-full spike IgG (median 1.23 S/Co, IQR: 0.61-1.73 vs 2.08 S/Co, IQR: 1.45-3.04, p=0.62) were slightly lower, but not significantly reduced when correcting for multiple testing following prime immunisation with ChAdOx compared to BNT (**Fig. 3B, Fig. S1**). Three weeks after boost immunisation, antibody responses in homologous BNT/BNT immunised participants was comparable to heterologous ChAdOx/BNT immunised participants (anti-S1 IgG median: 4.52 S/Co, IQR:3.92-5.10 vs 5.37 S/Co, IQR: 4.82-5.86, p=0.31) (**Fig. 3A**,**B, Fig. S1**).

**Fig. 3.**
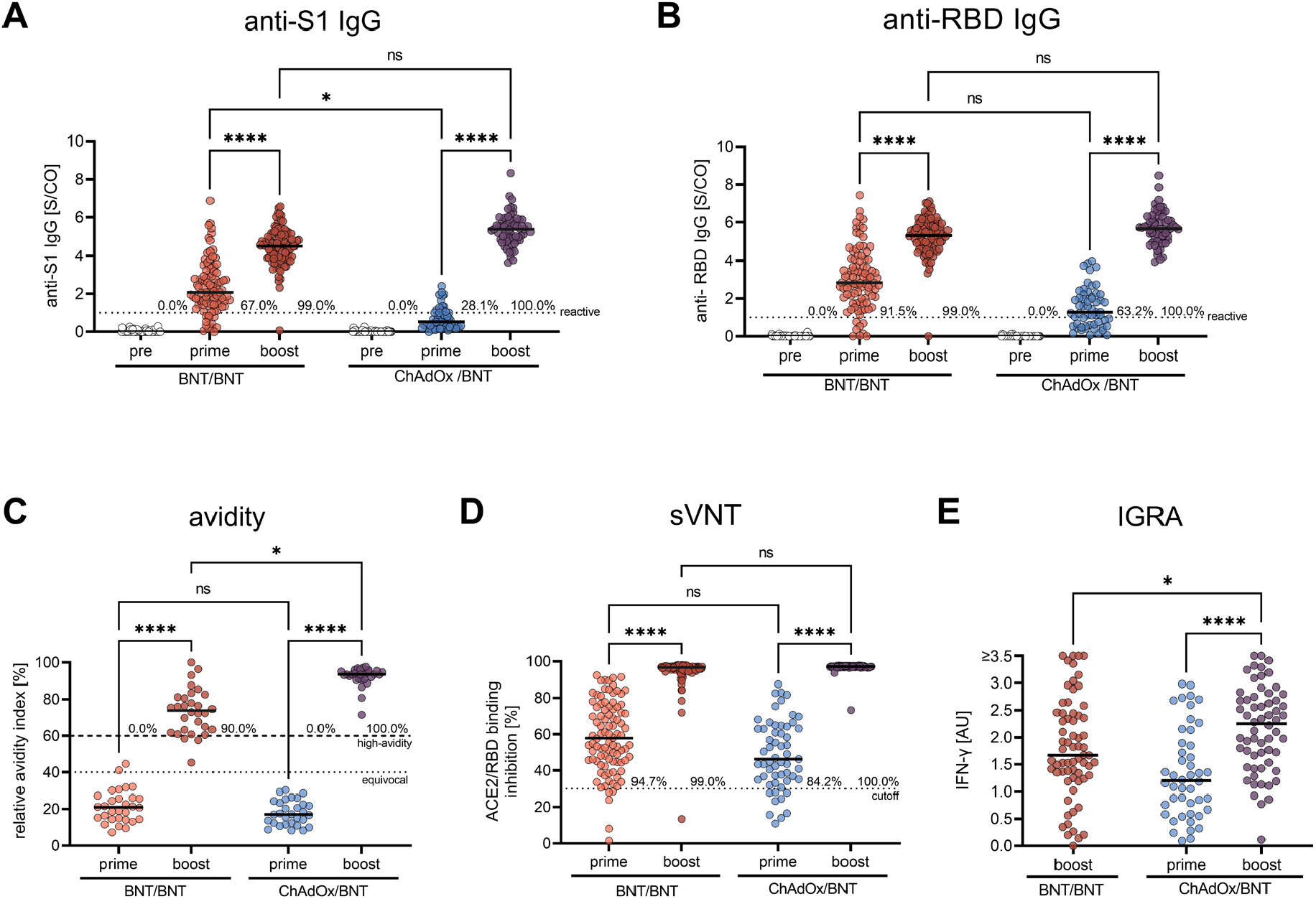
SARS-CoV-2 specific antibody- and T cell response after BNT or ChAdOx prime immunisations and homologous or heterologous booster vaccination. (**A**) Anti-S1 IgG and (**B**) anti-RBD IgG measured by SeraSpot Anti-SARS-CoV-2 IgG assay, (**C**) anti-S1 IgG avidity, and (**D**) neutralizing capacity measured by sVNT in serum of subjects who had received prime immunisation with BNT or ChAdOx, and homologous BNT/BNT or heterologous ChAdOx/BNT boost. (**E**) T cell reactivity in whole blood samples measured by IGRA. BNT: BNT162b2 / Comirnaty, ChAdOx: ChAdOx1-nCoV19 / Vaxzevria. IgG: Immunoglobulin G, S/CO: signal-to-cutoff ratio, sVNT: surrogate virus neutralization assay, ACE2: angiotensin-converting enzyme 2, RBD: SARS-CoV-2 receptor-binding domain, S1: SARS-CoV-2 Spike protein S1 domain, AU: arbitrary unit. Sampling time points: pre: pre-immune sample prior to first immunisation, prime: three weeks after first vaccination, boost: three to four weeks after boost vaccination. Dotted lines indicate the manufacturer’s pre-specified threshold, for anti-RBD IgG >1 S/Co, for sVNT >30% and for avidity 40-60%: borderline avidity, >60%; high avidity. Lines indicate the median. * = p<0.05; ** = p<0.01; **** = p<0.000; ns: not significant.

In addition to antibody levels, we measured serum antibody avidity. High avidity serum antibodies, defined as an antibody avidity index >60%, were not detected after prime immunisation with either BNT or ChAdOx (**Fig. 3C**). Three weeks after boost immunisation 27/30 (90.00%, 95%CI: 74.38-96.54) participants in the homologous BNT/BNT group and 30/30 (100.00, 95%CI: 94.08-100.00) in the heterologous ChAdOx/BNT immunised group exhibited high anti-S1 IgG avidity indices (**Fig. 3C**). Hence, maturation of IgG avidity following boost vaccination was observed with both regimens. The median relative avidity index was slightly higher after heterologous ChAdOx/BNT boost (93.50%, IQR: 91.10-95.41) compared to homologous BNT/BNT boost (73.86%, 95%CI: 62.99-81.55, p=0.04, **Fig. 3C**), which may also be due to the longer dosing interval in the heterologous boost group.

Neutralising antibodies were detected in 89/94 (94.68%, 95%CI: 88.15-97.01) participants receiving BNT and in 48/57 (84.21%, 95%CI:72.64-91.46) participants receiving ChAdOx prime vaccination (**Fig. 3D**). At week three after boost immunisation with BNT, neutralising antibody response rate had increased in both cohorts to 100/101 (99.01%, 95%CI: 94.60-99.95) after BNT/BNT boost and 61/61 (100.00%, 95%CI: 94.08-100.00) after heterologous ChAdOx/BNT boost (**Fig. 3D**). Surrogate virus neutralisation test (sVNT) titers were comparable after homologous and heterologous prime-boost immunisation (**Fig. 3D**).

Serological responses are most widely used to assess immunogenicity of vaccination, but T cell responses are another important marker of anti-SARS-CoV-2 immunity. The spike S1-specific T cell response was measured in 47 ChAdOx prime immunised, 60 ChAZOx/BNT boost immunised and 66 BNT/BNT boost immunised subjects by IFN-γ release (IGRA). Three weeks after ChAdOx prime immunisation, participants showed robust T cell responses (**Fig. 3E**). Notably, T cell reactivity was significantly higher after heterologous ChAdOx/BNT boost immunisation compared to homologous BNT/BNT boosting (1.67 AU, IQR: 1.29-2.45, vs. 2.25 AU, IQR: 1.57-2.73, p=0.0255) (**Fig. 3E**).

## Discussion

This observational cohort study involving 340 health care workers provides real-world data on reactogenicity and immunogenicity of homologous BNT/BNT immunisation compared to heterologous ChAdOx/BNT vaccination against COVID-19. Overall, both regimens were well-tolerated. We observed no major difference in reactogenicity between both prime-boost regimens. Overall, local reactions were frequently observed for all vaccines. Systemic reactions, including severe reactions, were most frequent following prime immunisation with ChAdOx, whereas reactogenicity of BNT/BNT and ChAdOx/BNT was comparable, with slightly decreased systemic reactions of the heterologous booster. We observed robust immunogenicity of both homologous and heterologous prime-boost regimens. Increased S1-reactive T cell responses as measured by IGRA, were increased three weeks after heterologous ChAdOx/BNT boost compared to BNT/BNT boost vaccination. Thus, heterologous ChAdOx/BNT immunisation with a vaccine interval of 10-12 weeks is well tolerated and highly immunogenic, comparable to homologous BNT/BNT vaccination.

### Strengths and limitations of this study

This is the first report of immunogenicity of heterologous ChAdOx/BNT compared to homologous BNT/BNT prime-boost vaccination. This is also the first report of real-world reactogenicity of ChAdOx/BNT vaccination with a 10-12 week vaccine interval, compared to BNT/BNT vaccination with a 3-week vaccine interval. Another strength is the longitudinal follow-up of up to 15 weeks after first immunisation. Data of this nature is urgently needed due to ongoing heterologous vaccinations in several countries.

Our study also has several potential limitations, as it is not a randomized controlled trial. Due to the current recommendations for heterologous ChAdOx/mRNA vaccination in persons <60 years of age, we were not able to recruit a matched cohort of homologous ChAdOx/ChAdOx vaccinated health care workers, since most of the study participants opted for the recommended heterologous booster. Hence, we cannot determine the exact effect of the heterologous BNT booster vaccine compared to ChAdOx homologous boosting alone. This is an interim analysis as the study is still recruiting and a comparison with homologous ChAdOx/ChAdOx vaccination may be possible with the next analysis. Here, we compared reactogenicity and immunogenicity of homologous BNT/BNT and heterologous ChAdOx/BNT vaccination. In addition to the different combinations of prime and boost vaccines, the interval between first and second vaccine was significantly different in the homologous (21 days) and heterologous vaccination group (71 days) (**Table 1**). Thus, it is unclear to which extent the observed differences may also be attributable to the extended vaccine interval in the heterologous vaccination group. The observed increased anti-S1 IgG avidity, for instance, is likely to be caused by the extended vaccination interval, since antibody affinity maturation increases over time.

### Comparison to other studies

We observed comparable reactogenicity of homologous BNT/BNT vaccination and heterologous ChAdOx/BNT vaccination, both of which were well-tolerated in our cohort with a 10-12 week dosing interval. This is in contrast with interim results of the Com-COV trial, which reported increased systemic vaccine reactions following heterologous ChAdOx/BNT vaccination, compared to homologous ChAdOx/ChAdOx and BNT/BNT regimens in a comparable sample size [10]. There are several differences in the study design (RCT vs. observational study), study population demographics, and vaccine interval that may explain this discrepancy. The median age in Com-CoV was 57 years (46% females), and 34 years (29-45 years, 60.18% females) in the present study (**Table 1**). The interval between first and second vaccination with either BNT or ChAdOx was 28 days in the Com-COV study, compared to 71 days reported here. We hypothesize that extending the vaccine interval to 10-12 weeks may limit the reactogenicity of heterologous ChAdOx/BNT vaccination.

Phase 1/2 studies have previously reported robust immunogenicity of homologous BNT and ChAdOx immunisations [21,22]. In contrast, immunogenicity of heterologous ChAdOx/BNT immunisation has not been previously reported. Our data indicate that both homologous and heterologous regimens induced high titers of high-affinity antibody responses and high T cell reactivity in healthy individuals. Whereas a slightly higher humoral response was noted after prime immunization with BNT compared to ChAdOx, we found no significant difference in antibody levels, or -neutralisation capacity at three weeks post homologous or heterologous booster vaccination, indicating that BNT booster immunisation induces strong humoral immune responses, even following weaker initial responses after ChAdOx prime immunisation. This is in line with previous studies reporting increased antibody responses in COVID-19 convalescents following a single dose of BNT, compared to seronegative persons receiving two doses of BNT [23]. Both vaccine regimens induced robust T cell responses, but, we observed slightly increased T cell reactivity after heterologous ChAdOx/BNT immunisation compared to the homologous BNT/BNT regimen, indicating that heterologous vaccination may increase immunogenicity.

### Policy implications

Heterologous prime-boost vaccination is currently recommended for individuals with ChAdOx prime immunisation in several countries, following reports of rare but serious adverse events associated with ChAdOx, particularly in younger women [7]. A heterologous boost with an mRNA vaccine (BNT or mRNA1273) with a vaccine interval of 12 weeks is currently recommended in Germany for persons under the age of 60 who have previously received one dose of ChAdOx [6]. Our study provides real-world evidence for the safety and immunogenicity of this vaccine regimen. Heterologous vaccination schedules might also alleviate logistical challenges and mitigate intermittent supply shortages of individual vaccines. In light of increasing occurrence of new virus variants carrying immune escape mutations, it will be important to determine whether heterologous vaccination regimens might enhance protection against infection and severe COVID-19. Further controlled studies are required to answer this question.

### Conclusions

In summary, this study provides evidence that heterologous ChAdOx/BNT immunisation with 10-12 week intervals, currently recommended in several countries, is well tolerated and equally immunogenic as homologous BNT/BNT vaccination, with evidence of enhanced T cell responses. Our data support further studies into the applicability of heterologous prime-boost vaccination strategies for COVID-19.

## Data Availability

Data is available upon reasonable request.

## Acknowledgements

The authors are grateful to all study participants at Charité - Universitätsmedizin Berlin for their participation. The authors also thank the entire staff of the Department for Occupational Medicine and the Charité Clinical Study Center (CSC) at Charité - Universitätsmedizin Berlin and the Berlin Institute of Health (BIH) for their support of the study.

This study was supported by the Forschungsnetzwerk der Universitätsmedizin zu Covid-19, COVIM - FKZ: 01KX2021 (to L.E.S., V.M.C., F.K., C.D., N.S.).

## Supplementary Material

**Figure S1:**
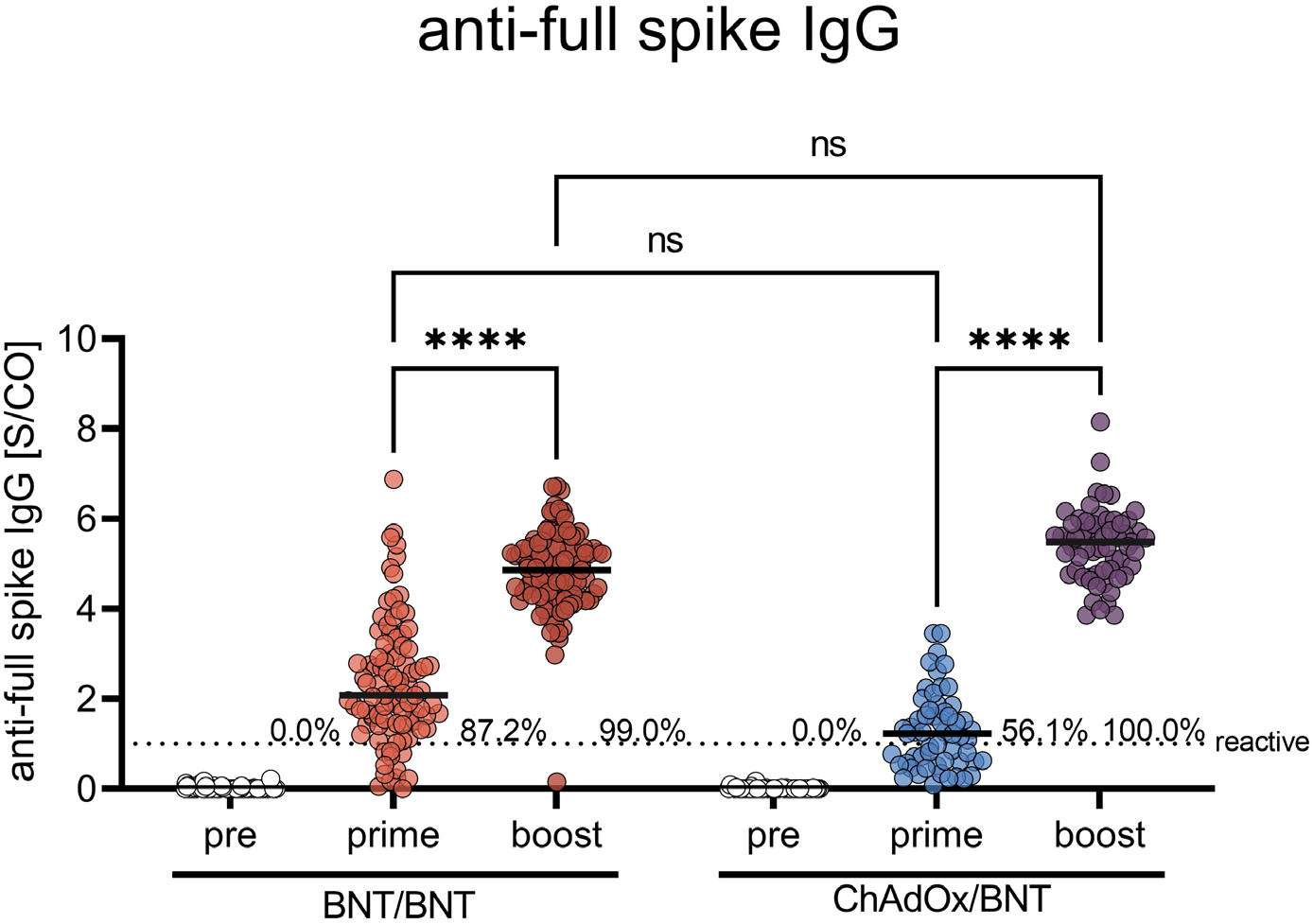
Serum anti-full spike IgG response after BNT or ChAdOx prime immunisations and homologous or heterologous booster vaccination. Anti-full spike-IgG in serum measured by SeraSpot Anti-SARS-CoV-2 IgG assay. BNT: BNT162b2 / Comirnaty; ChAdOx: ChAdOx1-nCoV19 / Vaxzevria; IgG: Immunoglobulin G; S/CO: signal-to-cutoff ratio. Sampling time points: pre: pre-immune sample prior to first immunisation; prime: three weeks after first vaccination; boost: three to four weeks after boost vaccination. The dotted line indicates the manufacturer’s pre-specified threshold (>1 S/Co). Lines indicate the median. **** = p<0.0001; ns: not significant.

**Table S1:** Local and systemic reactogenicity of BNT or ChAdOx prime immunisations and homologous or heterologous boosting until day 7 after vaccination. Proportion of participants reporting local and systemic reactions and intake of antipyretic medication per group (95% CI). 95% confidence intervals were calculated according to the Wilson and Brown method.

| **Local reaction** | **1<sup>st</sup> BNT** | **1<sup>st</sup> BNT / 2<sup>nd</sup> BNT** | **1<sup>st</sup> ChAdOx** | **1<sup>st</sup> ChAdOx / 2<sup>nd</sup> BNT** |
| --- | --- | --- | --- | --- |
| Local reaction (any severity) | 69,10<br>(61,97 - 75,43) | 74,21<br>(66,9 - 80,39) | 81,08<br>(74,02 - 86,57) | 84,85<br>(76,5 - 90,6) |
| Local reaction (only severe) | 1.68<br>(0.45 - 4.83) | 3.77<br>(1.74 - 7.99) | 3.38<br>(1.45 - 7.66) | 5.05<br>(2.18 - 11.28) |
| Pain at injection site (any severity) | 62.36<br>(55.05 - 69.15) | 69.81<br>(62.28 - 76.41) | 77.70<br>(70.34 - 83.66) | 79.80<br>(70.85 - 86.52) |
| Tenderness (any severity) | 68.54<br>(61.39 - 74.91) | 73.58<br>(66.23 - 79.82) | 80.41<br>(73.28 - 86.00) | 83.84<br>(75.35 - 89.80) |
| Swelling (any severity) | 3.93<br>(1.92 - 7.89) | 7.55<br>(4.37 - 12.73) | 3.38<br>(1.45 - 7.66) | 6.06<br>(2.81 - 12.60) |
| Redness (any severity) | 2.25<br>(0.88 - 5.63) | 1.89<br>(0.51 - 5.40) | 2.70<br>(1.06 - 6.74) | 6.06<br>(2.81 - 12.60) |
| **Systemic reaction** |  |  |  |  |
| Systemic reaction (any severity) | 38.76<br>(31.92 - 46.09) | 64.78<br>(57.09 - 71.78) | 86.49<br>(80.05 - 91.08) | 51.52<br>(41.80 - 61.12) |
| Systemic reaction (only severe) | 0.56<br>(0.03 - 3.11) | 6.29<br>(3.45 - 11.19) | 22.97<br>(16.93 - 30.38) | 2.02<br>(0.36 - 7.07) |
| Headache (any severity) | 17.42<br>(12.55 - 23.66) | 38.99<br>(31.76 - 46.75) | 75.00<br>(67.45 - 81.28) | 37.37<br>(28.48 - 47.21) |
| Fatigue (any severity) | 24.16<br>(18.46 - 30.95) | 47.80<br>(40.18 - 55.52) | 72.97<br>(65.30 - 79.48) | 37.37<br>(28.48 - 47.21) |
| Feverishness & chills (any severity) | 8.99<br>(5.61 - 14.1) | 19.50<br>(14.09 - 26.34) | 70.27<br>(62.47 - 77.05) | 18.18<br>(11.82 - 26.92) |
| Myalgia (any severity) | 20.79<br>(15.47 - 27.33) | 33.96<br>(27.06 - 41.62) | 54.73<br>(46.69 - 62.53) | 22.22<br>(15.16 - 31.36) |
| Arthralgia (any severity) | 3.93<br>(1.92 - 7.89) | 10.06<br>(6.29 - 15.72) | 29.73<br>(22.95 - 37.53) | 8.08<br>(4.15 - 15.14) |
| Nausea & vomiting (any severity) | 3.93<br>(1.92 - 7.89) | 4.40<br>(2.15 - 8.81) | 15.54<br>(10.58 - 22.24) | 6.06<br>(2.81 - 12.60) |
| Diarrhoea (any severity) | 3.93<br>(1.92 - 7.89) | 6.29<br>(3.45 - 11.19) | 7.43<br>(4.20 - 12.82) | 4.04<br>(1.58 - 9.93) |
| **Antipyretic medication** |  |  |  |  |
| Intake within first 24 hours | 9.55<br>(6.05 - 14.76) | 22.64<br>(16.83 - 29.75) | 72.97<br>(65.30 - 79.48) | 32.32<br>(23.92 - 42.05) |
| Prophylactic intake | 2.81<br>(1.21 - 6.41) | 3.14<br>(1.35 - 7.15) | 27.03<br>(20.52 - 34.70) | 4.04<br>(1.58 - 9.93) |

